# Tumor Spectrum and Temporal Cancer Trends in adult carriers of Li-Fraumeni syndrome: Implications for Personalized Screening Strategies in *TP53* R337H carriers

**DOI:** 10.1101/2024.06.07.24308605

**Authors:** Pedro A. F. Galante, Gabriela D. A. Guardia, Janina Pisani, Renata L. Sandoval, Mateus C. Barros-Filho, Ana Carolina Leite Vieira Costa Gifoni, Diogo F. C. Patrão, Patricia Ashton-Prolla, Vitor Fiorin de Vasconcellos, Claire Freycon, Arnold Levine, Pierre Hainaut, Maria Isabel Achatz

## Abstract

**Background:** Li-Fraumeni Syndrome (LFS) is a predisposition associated with early onset malignant tumors caused by germline pathogenic variants in the TP53 gene. Although rare worldwide, LFS is prevalent in Southern Brazil due to a founder pathogenic variant, c.1010G>A, p.Arg337His (R337H), discovered through its association with risk of childhood adrenal cortical carcinoma.

**Methods:** Here, we have analyzed tumor patterns, cancer risk, sex differences and temporal tumor trends in a cohort of 303 adult R337H carriers and we have compared them with those of 405 adult carriers of other *TP53* variants typical of LFS.

**Findings:** Compared to the latter, R337H carriers had a lower cumulative risk of developing cancer (54% for R337H vs 78% for other *TP53* variants at age 50 years). Female R337H carriers were at a higher risk than males (65% vs 30% at age 50 years) and had a higher risk of developing a second primary cancer, underscoring a strong sex bias not observed in carriers of other variants. The most common cancers were breast (75%), soft tissue sarcoma (9%) and lung cancer (8%) in females, and soft tissue sarcoma (30%), prostate (13%) and lung cancer (12%) in males. Common second malignancies were breast cancer in females and lung cancer in males.

**Interpretations:** Overall, this study confirms that R337H is associated with a lifetime risk of multiple cancers of the LFS spectrum but with incomplete penetrance, particularly in males. Our findings suggest that R337H carriers would benefit from adapted lifetime and surveillance strategies for risk reduction.

**Funding:** São Paulo Research Foundation and Hospital Sírio-Libanês.

## INTRODUCTION

*TP53* is a multi-functional gene commonly mutated in human tumors, with more than 50% of cancers carrying loss-of-function somatic mutations in *TP53* ^1,2^. *De novo* germline pathogenic (P) or likely pathogenic (LP) *TP53* variants are thought to occur at a frequency of about 1:5000 births and carriers are associated with a lifetime predisposition of developing multiple primary tumors known as Li-Fraumeni Syndrome (LFS) ^3,4^. The LFS cancer spectrum is characterized by early life tumors including adrenocortical carcinoma, premenopausal breast cancer, bone and soft tissue sarcomas, brain tumors and hematopoietic malignancies , among others ^5^.

In 2001, Ribeiro and colleagues reported that an atypical *TP53* variant, c.1010G>A; p.Arg337His (R337H), was frequently found in the germline of children with adrenocortical carcinoma in Southern Brazil ^6^. In subsequent years, a high number of children and adult carriers of this variant were detected in the same area, presenting complete or partial LFS traits ^7^. R337H differs from most *TP53* P/LP variants by its location in exon 10, encoding the oligomerization domain of the p53 protein, whereas the majority of variants causing LFS occurs in exons 4-9 (encoding the DNA-binding domain). R337H variant is carried by a founder haplotype of Lusitanian/Hispanic origin, estimated to be present in about 0.3% of the population of Southern Brazil ^8–12^. Over the past 15 years, the magnitude of cancer risk caused by R337H has been a matter of debate, some carriers presenting with full LFS traits and many others with attenuated LFS features or remaining apparently cancer-free over lifetime. Given its high prevalence, R337H is a significant cancer risk in the Brazilian population. Therefore, detailed information on lifetime cancer risk, tumor spectrum and risk of multiple primary tumors is crucial for developing awareness and appropriate screening and risk reduction strategies ^13^. Of note, with the development of genome sequencing as a basis for molecular diagnosis, R337H represents a frequent finding in adults, but current knowledge of tumor profiles in R337H adult carriers is scarce and limited to studies with few samples or individuals ^12^.

Here, we report on the Brazilian Li-Fraumeni Syndrome Study (BLiSS), the largest cohort of Brazilian LFS adults available to date. We provide a detailed assessment of tumor spectrum, lifetime cancer risk, temporal tumor trends and sex differences associated with R337H and we compare them with those associated with other P/LP variants found in Brazilians or in carriers from other origins. We show that R337H entails a significant lifetime risk of multiple cancers, which recapitulates many, but not all traits of LFS, with striking differences between males and females. Overall, these results suggest that tailored screening and risk reduction strategies could be developed and adapted to the needs of R337H carriers.

## METHODS

### Brazilian Li-Fraumeni Syndrome Study (BLiSS) cohort

We retrospectively investigated tumor spectrum, clinical and demographic status in a cohort of 708 adults with germline TP53 P/LP variants. P/LP variant status was based on ClinGen TP53 Expert Panel Specifications to the ACMG/AMP Variant Interpretation Guidelines for TP53 (Version 1.3.0, https://cspec.genome.network/cspec/ui/svi/doc/GN009). The cohort was constituted from two datasets: (1) BLiSS, comprising 345 Brazilian adult heterozygous carriers (age range 18-82 years, median 42 years), including 303 carriers (88%, 160 families) of *TP53* c.1010G>A (R337H) and 42 carriers (12%, 29 families) of other *TP53* P/LP variants (non-R337H carriers); (2) 363 adult carriers of non-R337H variants (age range 18-90, median 38 years) from diverse geographic areas, extracted from the NCI *TP53* database (version R20; https://tp53.isb-cgc.org/), a resource that compiles annotations on *TP53* carriers from the literature (**Supplementary Table S1**).

Individuals recruited into BLISS were detected between 2018 and 2022 in a high-cancer risk familial clinics at Hospital Sírio-Libanês (Sao Paulo, Brazil). All subjects tested positive for a germline P/LP *TP53* variant either by conventional targeted gene sequencing or by multigene next generation sequencing panels. All clinical and personal data were de-identified before data sharing and analyses. This study was approved by the institutional research ethics committees in all centers (approval at the coordinating center, Hospital Sírio-Libanês #3.830.276). All patients signed a written informed consent. Data from the NCI TP53 dataset are fully anonymous and open-source.

### Statistical analyses

Data processing, statistical analyses and figures were produced in the R software (version 4.0.3), using the following R packages: ggplot2 (v3.3.6), ggpubr (v0.4.0), survival (v3.2.13), survminer (v0.4.9), tidyverse (v1.3.1), trackViewer(v1.24.2) and webr (v0.1.5). Figure colors and aesthetic edition were performed using the Inkscape software (https://inkscape.org/). Sankey diagrams were created using the SankeyMATIC web tool (https://sankeymatic.com/). Statistical comparisons between patient groups were performed using Wilcoxon signed-rank tests and significance of Kaplan Meier curves were assessed by log-rank tests. All statistical tests were considered significant at p-value < 0.05, and, if applicable, adjusted to multiple test comparisons using false discovery rate.

## RESULTS

### Study cohort

The goal of this study was to rigorously compare cancer phenotypes and lifetime risk in Brazilian adult R337H carriers with those of adult carriers of other P/LP TP53 variants (non-R337H), either from Brazil or from other regions of the world. R337H carriers were identified within the BLiSS study and the comparison group was constructed by lumping non-R337H carriers from BLiSS with a subset of carriers with similar age range and median age as BLISS carriers, taken from the public NCI TP53 dataset (see methods). Overall, we compiled data on a total of 708 adults with P/LP variants, including 345 subjects from BLiSS registry and 363 from the NCI *TP53* database (https://tp53.isb-cgc.org/), **Figure 1A**. Of these 708 carriers, 303 (160 families) carried the founder R337H variant and 405 carried non-R337H P/LP variants (172 families), including 42 Brazilian carriers (29 families) (**Figure 1A**). **Figure 1B** shows the occurrence and position of these variants with respect to p53 domains. Aside from R337H (303 occurrences), which is located in the oligomerization domain, a total of 46 different other variants were identified, mostly located within the DNA-binding domain. These variants included the missense variants M133T (c.398T>C, n=22 carriers), R175H (c.524G>A, n=21), T125= (c.375G>A, n=19), R248W (c.742C>T, n=19), G245S (c.733G>A, n=18), R213Q (c.638G>A, n=16) and R273H (c.818G>A, n= 15), as well as the silent variant T125= (c.375G>A, n=19), which has been shown to inactivate p53 by disrupting gene splicing and preventing p53 mRNA synthesis ^14^. This dataset represents the largest cohort to date of adult carriers of P/LP TP53 variants, including R337H carriers.

**Figure 1.**
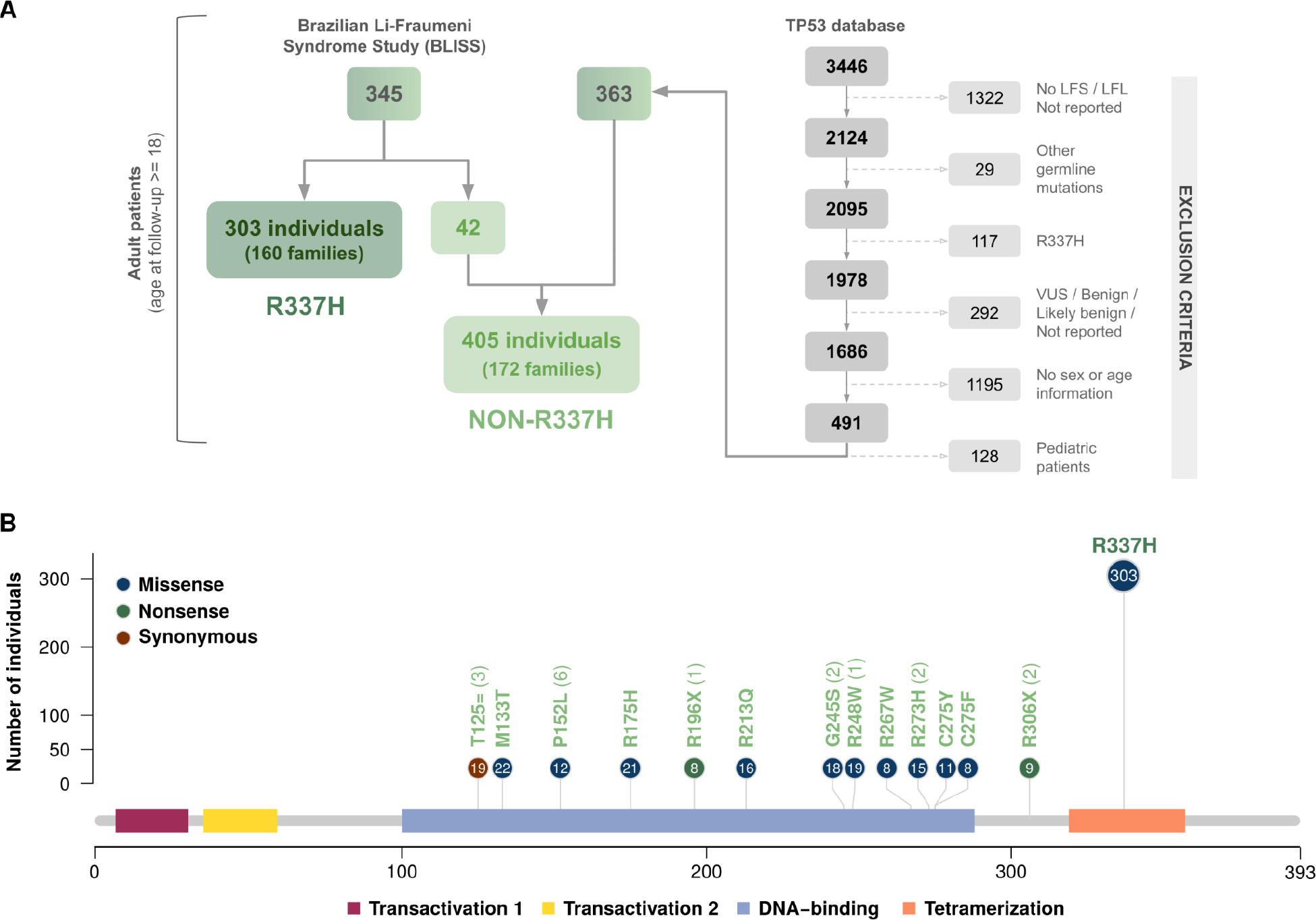
Overview of R337H and non-R337H study cohorts. A) Study flowchart showing the selected R337H (n=303) and non-R337H (n=405) individuals. B) Frequency of *TP53* variants included in our study and affected *TP53* domains. Variations found in at least 1% of individuals are shown. Number of non-R337H carriers only from the BLiSS cohort is shown in parenthesis when appropriate.

### Cancer occurrence and patterns in R337H carriers

R337H carriers presented with significantly less cancer diagnosis than non-R337H carriers(log-rank test p-value < 0.0001, **Figure 2A**). At age 50 years, 54% (162/303) of R337H carriers had at least one cancer diagnosis, compared to 78% (316/405) of non-R337H carriers. Noteworthy, by the age of 65, 78% R337H and 81% non-R337H carriers had at least one cancer diagnosis, respectively, suggesting that R337H tended to develop cancers at later ages than non-R337H. Among R337H carriers, 242 tumors were diagnosed in 162 carriers, while 141 carriers (46.5%) did not present with a cancer diagnosis. The most common diagnoses were breast cancer (n=114, 47.1%), soft-tissue sarcoma (n=37, 15.3%) and lung cancer (n=28, 11.6%), comprising about 75% of all diagnoses in R337H carriers (**Figure 2B**). Among non-R337H carriers, the most frequent diagnoses were breast cancer (n=145, 33.6%), brain cancer (n=48, 11.1%) and soft tissue sarcoma (n=35, 8.1%), comprising 52.8% of all tumors diagnosed in this group, whereas 67 carriers (16.5%) were cancer-free. Overall, 4 cancer types were proportionally more represented in R337H than non-R337H carriers: breast cancer (47.1% vs 33.6%, Fisher’s test p-value < 0.001), soft tissue sarcoma (15.3% vs 8.1%, Fisher’s test p-value = 0.006), lung cancer (11.6% vs 5.8%, Fisher’s test p-value = 0.01), and thyroid cancer (2.9% vs 0.7%, Fisher’s test p-value = 0.04). In contrast, brain cancer was more represented in non-R337H carriers (Fisher’s test p-value < 0.001), as well as bone sarcoma (Fisher’s test p-value < 0.0001), a cancer that represented 5.1% (n=22) of diagnoses in non-R337H carriers but was not observed in R337H carriers (Figure 2B).

**Figure 2.**
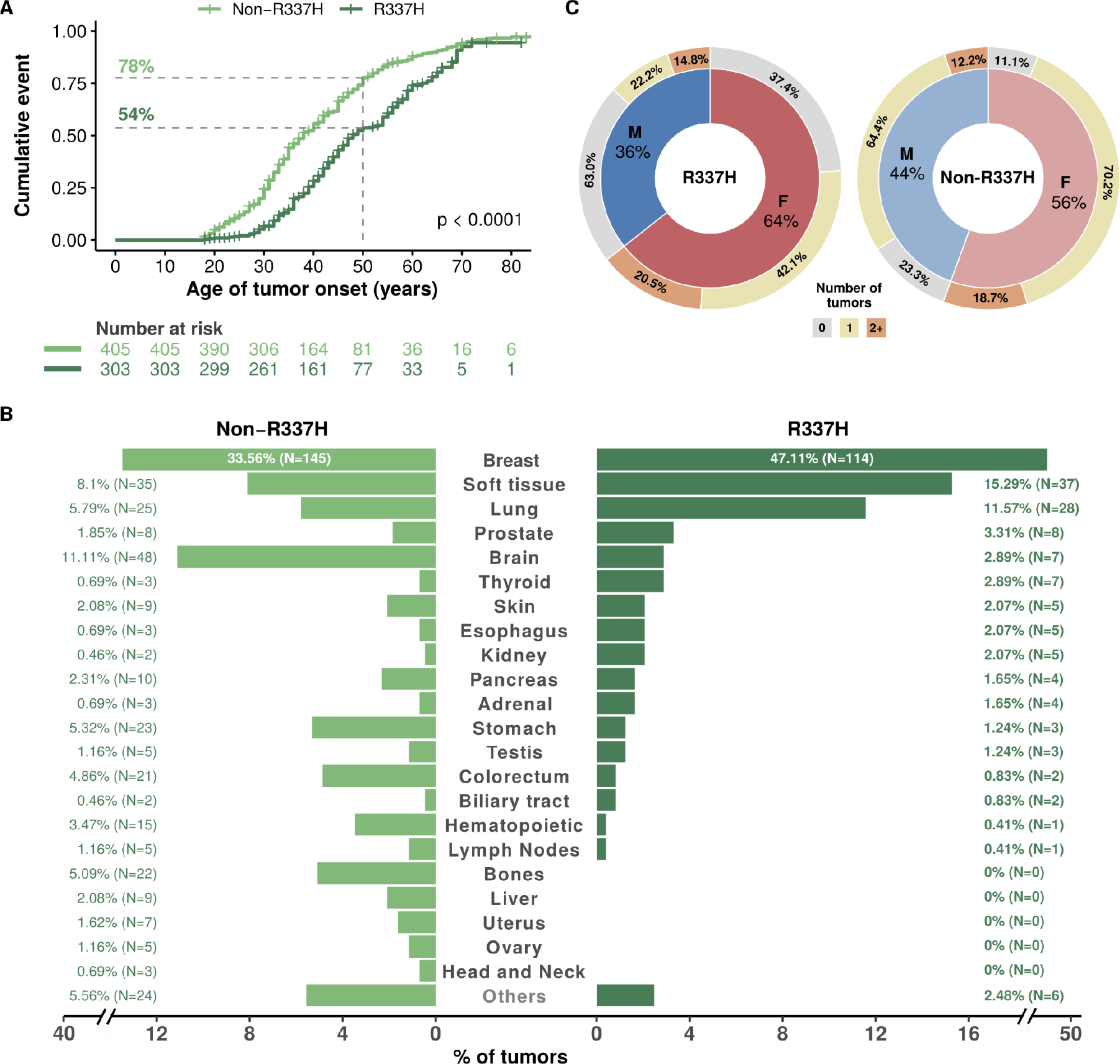
**R337H carriers have less and distinct cancer patterns than other non-R337H carriers**. A) Kaplan-Meier curves showing the cumulative risks of R337H and non-R337H carriers of developing a (first) primary tumor; B) Cancer patterns in R337H and non-R337H carriers; C) Proportions of male (M) and female (F) carriers without reported tumors, diagnosed with 1 primary tumor or with 2 or more primary tumors in R337H and non-R337H cohorts.

Overall, these results show that R337H carriers retained a high risk of cancer during adult life. Although they presented significantly less cancers than age-matched non-R337H carriers, R337H carriers had a high lifetime risk of breast cancer and soft tissue sarcoma, two cancers typical of the Li-Fraumeni cancer spectrum. However, they differed from non-R337H carriers by a lower proportion of brain cancers and by the conspicuous absence of bone sarcoma, a type of cancer considered as a signature of the LFS spectrum.

### Differences between R337H male and female carriers

Striking differences between R337H and non-R337H carriers were even evident after stratifying by sex. Among female R337H carriers (median age: 44 years), 122/195 (62.6%) were diagnosed with at least one cancer, while 200/225 (88.9%) of female non-R337H carriers (median age: 38 years) had one or more primary tumors (Fisher’s exact test; p-value < 0.0001, **Figure 2C and Supplementary** Figure 1). For males, only 40/108 (37.0%) of R337H carriers presented a cancer diagnosis while in male non-R337H carriers, 138/180 (76.7%) had at least one tumor diagnosis (Fisher’s exact test; p-value < 0.0001). This difference is remarkable given the median age at follow-up, which is higher in R337H (44 years) than in non-R337H (38 years) (Wilcoxon test p-value < 0.05; **Figure 2C, Supplementary** Figure 1).

To better characterize these differences, we estimated the cumulative risks of developing a first primary cancer in R337H and non-R337H carriers stratified by sex. Male carriers were significantly older at first diagnosis than females in both R337H (median ages: 46 and 41 years, respectively) and non-R337H groups (median ages: 38 and 35 years, respectively) (**Figure 3A**). Among R337H carriers, both males and females showed significantly lower lifetime risks of developing cancer than non-R337H counterparts (**Figure 3B**). While among non-R337H carriers, the cumulative cancer risk at 50 years was 82% for females and 72% for males, female R337H carriers had a 2 times greater cumulative risk of developing cancer than males at age 50 years (65% vs 30%, pairwise log-rank tests; FDR adjusted p-values: < 0.0001) (**Figure 3B)**.

**Figure 3.**
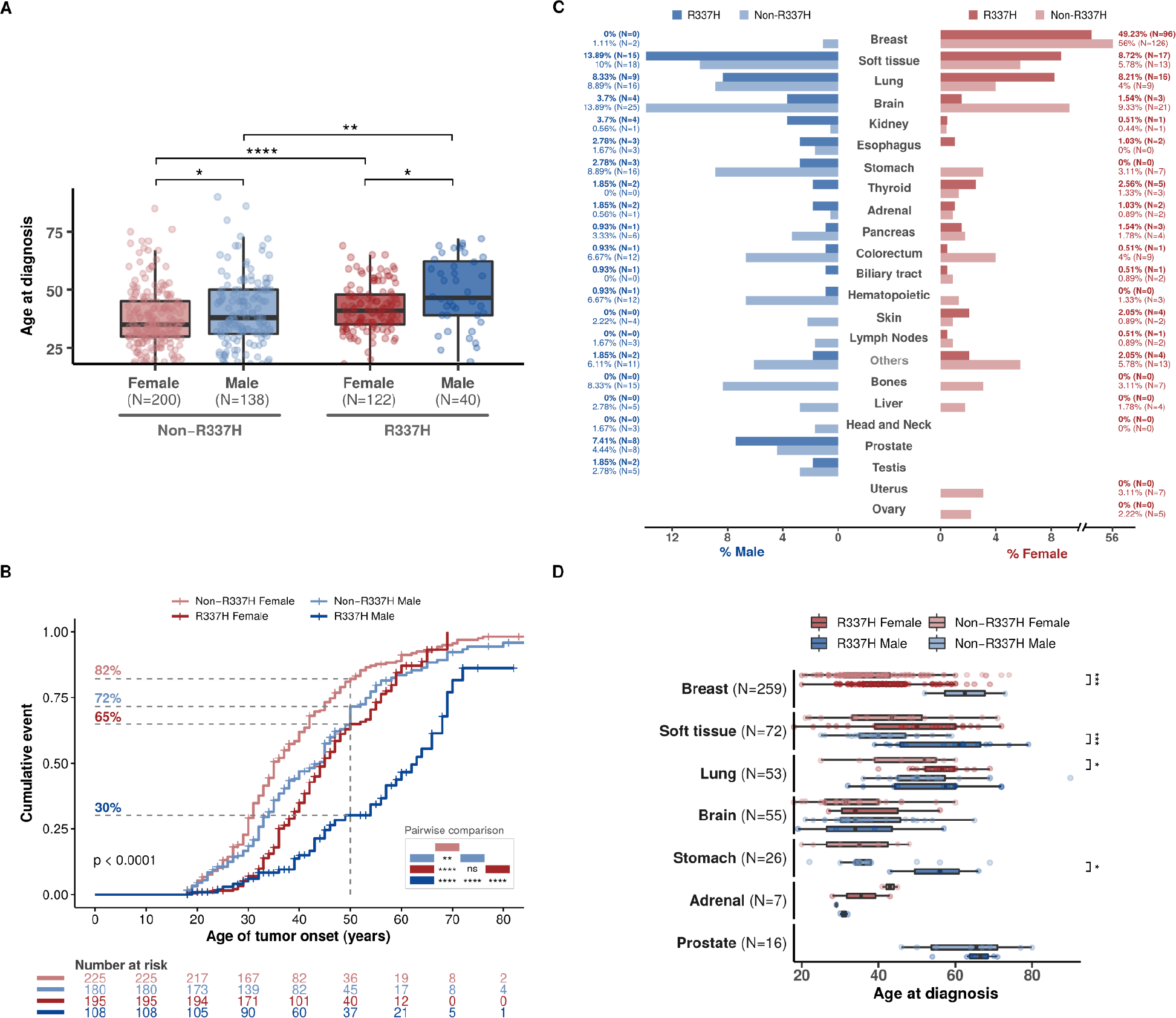
Cancer risk is lower among males and females carrying the R337H variant than non-R337H carriers, but females with R337H are at a higher lifetime cancer risk than males with the R337H variant. A) Ages of male and female individuals at first tumor diagnosis (Wilcoxon tests; p-values: * < 0.05, ** < 0.01, **** < 0.0001); B) Kaplan-Meier curves showing the cumulative risks of male and female individuals (R337H and non-R337H carriers) of developing a first tumor (pairwise log-rank tests; FDR adjusted p-values: **** < 0.0001, ** < 0.01, ns “not significant”); C) Tumor profile in male and female individuals carrying *TP53* R337H or non-R337H variants; D) Temporal patterns of tumor occurrence in male and female individuals (Wilcoxon tests; p-values: * < 0.05, *** < 0.001).

Sex-specific differences were also noted in the tumor spectrum (**Figure 3C**). Among females, lung cancer (8.2%), thyroid (2.6%) and skin tumors (2.0%) were the most common diagnoses in R337H carriers, but were proportionally rarer in non-R337H (4%, 1.3% and 0.9%, respectively). Conversely, brain cancers (9.3%) and colorectal cancers (4%) were more frequent in non-R337H than in R337H female carriers (1.5% and 0.5%, respectively) (Fisher test p-values 0.0005 and 0.02, respectively). Among males, R337H carriers presented more frequently with prostate cancer (7.4%) than non-R337H carriers (4.4%), whereas non-R337H had a greater proportion of brain (13.9%) and stomach cancers (8.9%) than R337H carriers (3.7% and 2.8%, respectively) (Fisher’s exact test p-values = 0.005 and 0.05, respectively). Of note, two cases of male breast cancers were detected among R337H carriers, compared to none in non-R337H carriers.

Next, we analyzed the temporal patterns of specific cancer in both sexes (**Figure 3D**). Both breast and lung cancers were diagnosed at later ages in female R337H carriers (median ages: 41 and 56.5 years) in comparison to non-R337H (median ages: 35 and 52 years) (Wilcoxon test p-values < 0.001 and <0.05, respectively). When considering male carriers, soft-tissue sarcomas and stomach cancers also showed delayed occurrence in R337H carriers (median ages: 61 and 56 years) compared to non-R337H carriers (median ages: 40 and 35 years) (Wilcoxon test p-values < 0.001 and <0.05, respectively). In contrast, prostate tumors tended to occur at similar median ages (>60 years) in R337H and non-R337H male carriers.

These observations show that cancer risk patterns differ between R337H and non-R337H adult carriers. For both sexes, R337H carriers had a lower lifetime risk of cancer, developing at later ages than non-R337H carriers. Furthermore, tumor patterns in R337H, while dominated by diagnoses typical of the LFS spectrum (breast cancer, soft tissue sarcoma) showed significant differences with non-R337H carriers, with a lower proportion of brain cancers and bone sarcoma and a higher proportion of lung and prostate cancers. Although these two cancers have been described as part of the LF spectrum ^3^, their detection in R337H carriers may phenocopy cancers that are common in the Brazilian population and should therefore be interpreted with caution. On the other hand, the low proportion of brain and bone cancers suggests that R337H carriers do not completely recapitulate the LF tumor spectrum. Furthermore, the sex bias observed in R337H but not in non-R337H carriers, suggests that exogenous/physiological factors associated with sex may modulate R337H penetrance and lifetime risk. The small number of patients analyzed is however a limitation and larger cohorts will be needed to confirm these differences.

### Lifetime risk and trajectories of multiple cancers

We investigated the occurrence of multiple malignancies in R337H carriers among 122 females and 40 males who were affected by a first primary cancer during adulthood. Overall, 40 females (32.8%) and 16 males (40.0%) had more than one tumor diagnosed, respectively (**Supplementary Table S1**). Both males and females with the R337H variant developed second tumors at later ages than non-R337H carriers Wilcoxon test p-values: < 0.05 and < 0.01, respectively), **Figure 4A**. Among R337H carriers, females appeared to be twice as likely than males to develop a second tumor by 50 years of age (32% vs 16%; log-rank test p-value < 0.01), **Figure 4B**. No such statistical differences could be observed among non-R337H carriers. Among R337H carriers, males had second primary malignancies at later ages than females (median ages: 61 and 47 years, respectively; Wilcoxon test p-value < 0.05), **Figure 4A**. These second malignancies were diagnosed on average 4 and 6 years after the first tumor in female and male carriers, respectively (**Supplementary** Figure 2).

**Figure 4.**
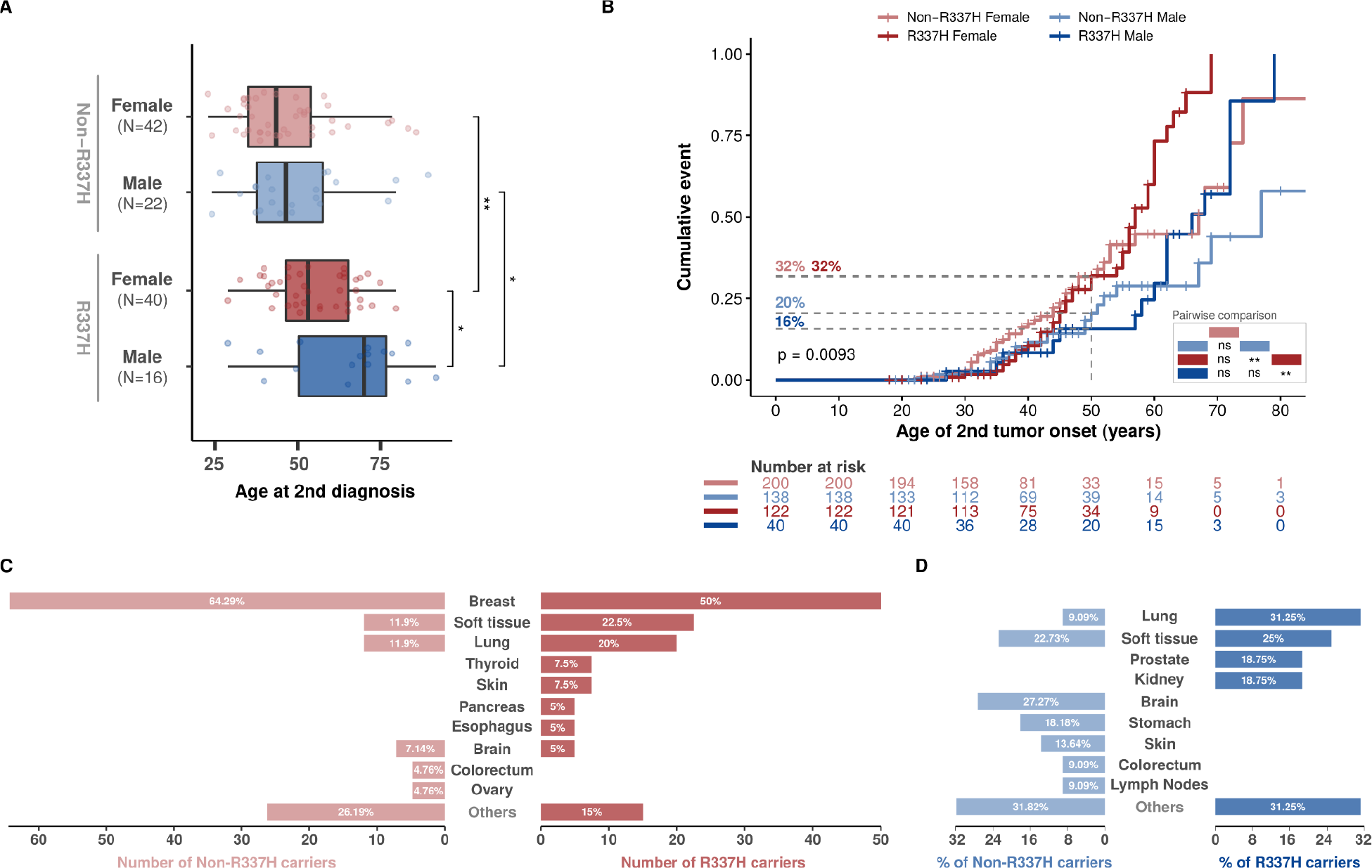
R337H females have a higher lifetime risk of developing a second tumor than R337H males. A) Ages of male and female individuals at second tumor diagnosis (Wilcoxon tests; p-values: * < 0.05, ** < 0.01); B) Kaplan-Meier curves showing the cumulative risks of male and female individuals (R337H and non-R337H carriers) of developing a second tumor (pairwise log-rank tests; FDR adjusted p-values: ** < 0.01, ns “not significant”); C-D) Proportions of additional tumors diagnosed in R337H and non-R337H C) female and D) male carriers (from second to last diagnosis).

Breast cancer was the most frequent second diagnosis among female carriers of any variant, affecting 50.0% (20/40) and 64.3% (27/42) of R337H and non-R337H carriers, respectively, **Figure 4C (Supplementary Table 2)**. Other frequent tumors included soft-tissue sarcoma (R337H: 22.5%, non-R337H: 11.9%) and lung cancer (R337H: 20.0%, non-R337H: 11.9%). Among male R337H carriers, the spectrum of second malignancies comprised predominantly lung cancer (31.2%), soft-tissue sarcoma (25.0%), prostate (18.7%) and kidney cancer (18.7%).

In non-R337H male carriers, the most frequent second tumors were brain cancers (27.3%), soft-tissue sarcomas (22.7%) and stomach cancers (18.2%), **Figure 4D**. Taken together, these results indicate that R337H female carriers are at a noticeably higher lifetime risk of developing multiple primary tumors and with earlier cancer occurrence patterns.

We finally examine the sequence in which multiple cancer may develop over adult lifetime in R337H and non-R337H carriers (**Figure 5**). In females, breast cancer was the most frequently first (R337H: 72.5%, non-R337H: 52.4%), second (R337H: 42.5%, non-R337H: 54.8%), and third (R337H: 33.3%, non-R337H: 50.0%) diagnosis, not only in patients who presented with first diagnosis of breast cancer but also in patients with any other first malignancy (**Figure 5A**). Besides breast cancer, female R337H carriers tended to develop lung (15.0%), soft-tissue sarcoma (10.0%), and thyroid cancer (7.5%) as their most common second diagnoses. Among males, the most frequent second diagnoses in R337H carriers were lung cancer (31%), kidney cancer (19%), and soft-tissue sarcoma (12%), whereas prostate cancer was the most common third diagnosis (50%) (**Figure 5B**). In contrast, in non-R337H male carriers, the most frequent second diagnoses were soft-tissue sarcoma (23%), brain cancer (23%), and gastric cancer (14%).

**Figure 5:**
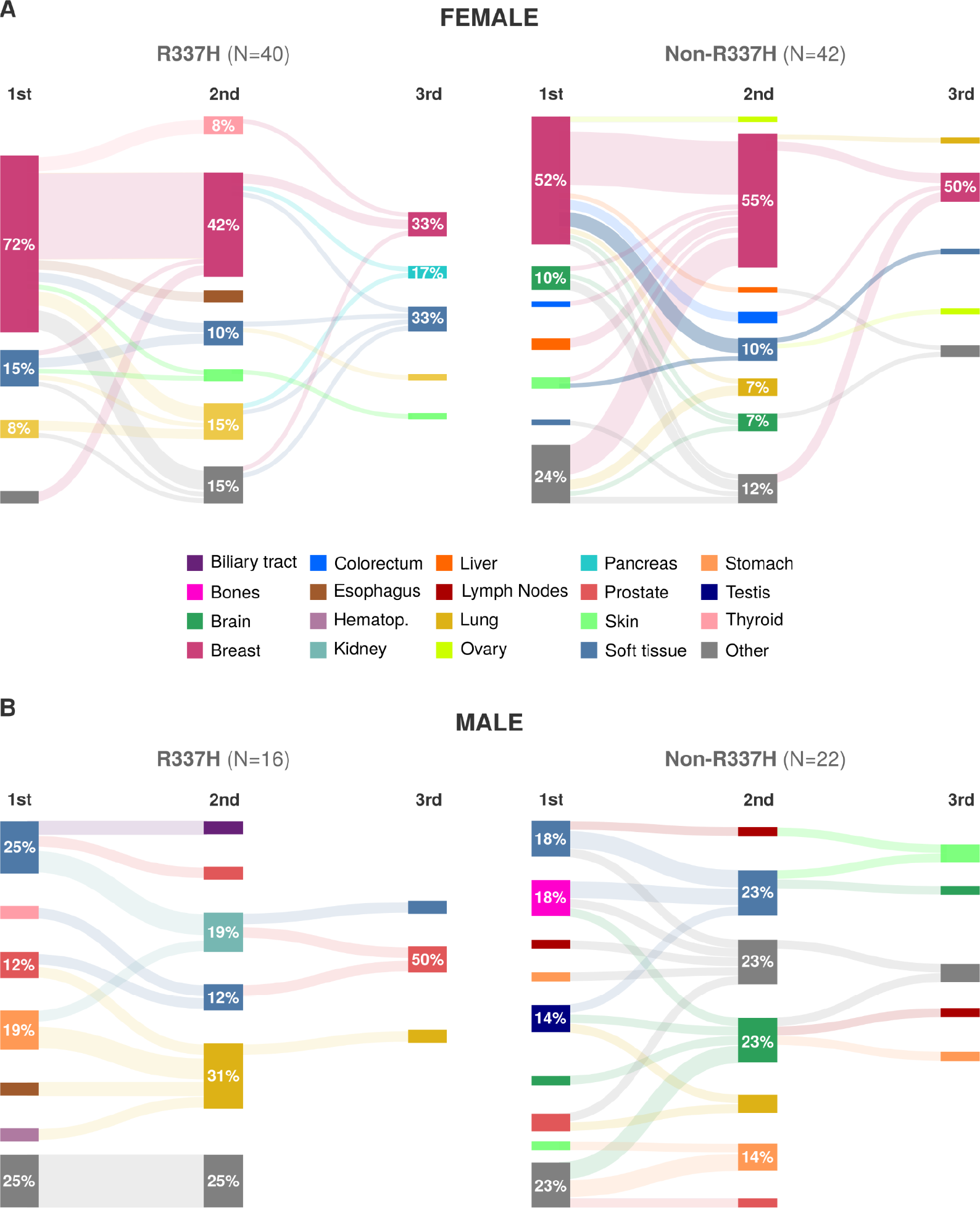
Distinct spectrum of second and third primary tumors in adult R337H carriers and non-carriers. Sankey diagrams show the patterns of occurrence of the first, second and third tumor types diagnosed in A) female and B) male individuals carrying the *TP53* R337H and non-R337H variants.

Overall, these observations support that R337H carriers have a significant lifetime risk of multiple cancers, with patterns of occurrence that differ from those of non-R337H carriers. Lung cancer, in particular, appeared to be a common second diagnosis in both male (31%) and female (15%) R337H carriers (compared to 9% and 7%, respectively, in non-R337H carriers).

## DISCUSSION

This study is the first systematic and comparative analysis of the risk and patterns of cancer in adult carriers of the TP53 R337H Brazilian founder allele versus carriers of other P/LP TP53 variants. It compiles and employs the largest database of adult carriers assembled to date, including 303 Brazilian R337H carriers from 160 families, compared with 42 carriers of non-R337H P/LP variants from 29 Brazilian families and 363 carriers of P/LP variants from 172 non-Brazilian families of mostly Caucasian background (see methods). Results clearly show that adult carriers of R337H allele have a lifetime pattern of risk that matches the Li-Fraumeni spectrum, characterized by excess risk of breast cancer in females and soft tissue sarcoma in both sexes. However, when compared to other P/LP variants, risk patterns in R337H carriers only partially recapitulates the LFS spectrum. The main traits that characterize adult R337 carriers versus other variants carriers (summarized in **Figure 6A**) are : (1) low prevalence, if not absence, of typical LF tumors such as brain cancers and osteosarcomas; (2) high prevalence of some cancers not considered as typical of the LF spectrum (renal cancer, thyroid cancers); (3) frequent occurrence of lung and prostate cancers, in particular as second diagnoses in males; (4) strong sex-related difference in lifetime risk, females having a risk almost twice higher than males. Overall, our observations further support that the cancer risk caused by the R337H variant is heterogeneous and dependent upon specific structural, functional and context-dependent variables that cause its particular tumor phenotype. Given the high population prevalence of R337H, our results will raise awareness of cancer risk in this adult population and will help in designing appropriate strategies for carrier surveillance and early diagnosis (**Figure 6B**).

**Figure 6.**
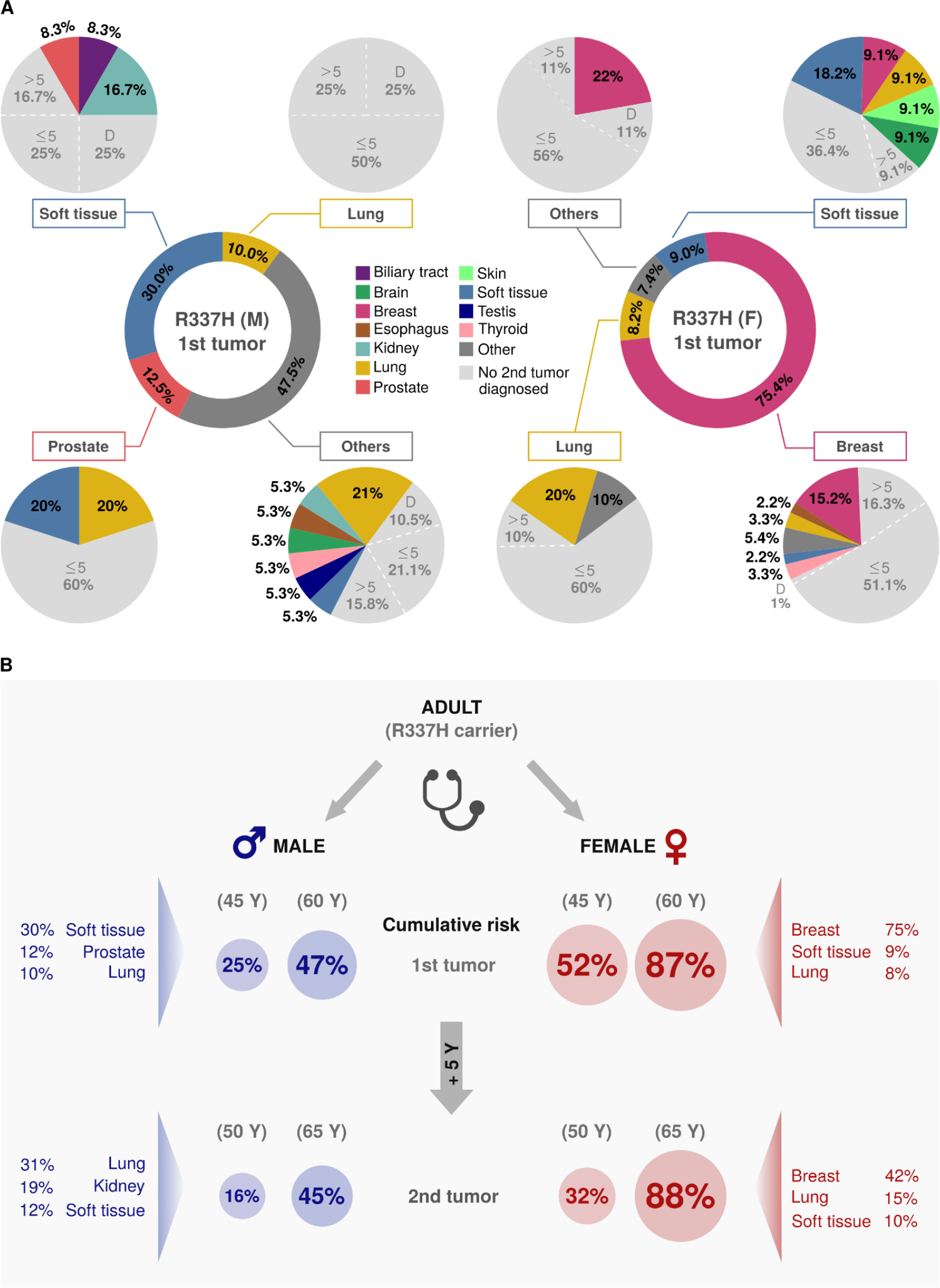
Our findings may guide clinicians in better screening R337H male and female adult carriers for the early detection of tumors. A) Pie/donut charts showing percentages of first (central pie charts) and second (outer pie charts) tumor types diagnosed in R337H carriers. Carriers with no second malignancies (in gray) were further stratified into: dead (D), alive for up to 5 years of follow-up (≤5) or more than 5 years (>5) ; B) Schematic representation of a decision tree to support clinical decisions on screening of R337H adult carriers based on our findings.

Any study of the variables that influence a genetic predisposition to develop cancer must take into account and explore those variables that impact the phenotypes of that cancer. Clearly, different TP53 variant alleles can influence cancer phenotypes as a consequence of their structural and functional impact on the p53 protein. In particular, TP53 variants occurring in different domains of the p53 protein may exert tissue specific effects. For example, somatic mutations in the p53 proline domain most often occur in skin cancers ^15^, suggesting enhanced effects of such variants in skin cells. The R337H variant is atypical among LF predisposing variants in its location in the p53 protein structure (tetramerization domain, whereas 80% of missense LFS variants fall in the DNA-binding domain) and its biochemical characteristics (uniquely sensitive to pH, enabling normal p53 oligomer assembly at neutral cell pH but preventing it at slightly elevated pH) ^16^. In vitro studies and mouse models have shown that the p.R337H protein can retain at least a degree of wild-type p53 transcriptional activities ^17^. Based on these observations, the R337H variant has been qualified as hypomorphic, potentially causing an attenuated phenotype compared to common P/LP variants. However, the present study suggests that this suspected hypomorphic effect does not equally affect LF features at all ages and in all tissues of R337H carriers.

An analysis of the cumulative risk of developing cancer at age 50 years (R337H=54% vs non-R337H=78%, p=< 0.0001) demonstrates that over the first 50 years of life, R337H carries a lower risk than other P/LP, but by age 65 years, these percentages are of 78% vs 81%, respectively, indicating that the rates of cancer occurrence strongly increases from 50-65 years of age in R337H carriers. Cancer patterns in this age group include postmenopausal Breast cancer, Sarcomas (in particular Leiomyosarcoma) as well as Lung or Prostate cancers. Thus age appears to be an important variable, with different TP53 alleles having different rates of cancer development as a function of age. Such selective effect of age on cancer risk has also been observed in children and adolescent carriers of R337H, who frequently develop Adrenal Cortical Carcinoma but show much fewer osteogenic sarcomas, choroid plexus papilloma/carcinomas, Rhabdomyosarcomas or Medulloblastomas than carriers of other P/LP variants ^7,18^. Moreover, the present analysis suggests that when young adult R337H carriers develop Glioblastoma, they enjoy a better survival under treatment than carriers of other P/LP variants. These observations clearly stress that the effect of the R337H variant is far from uniform across tissues and age and suggest that the risk of developing a particular cancer with different mutant TP53 alleles is largely tissue specific.

Indeed, the levels of p53 activity required to suppress cancer may differ from one tissue (or cell type) to another. Thus, a relatively modest attenuation of p53 activity caused by the R337H variant may be sufficient to a high risk of cancer in some tissues (breast, connective tissues) but not in others (bone, for example). The reasons underlying these cell or tissue differences in the sensitivity to p53 activity dosage are far from being understood.

Among other factors that may influence phenotypic variations in cancer predisposition, the genetic ancestry of the patient and the chromosomal context surrounding the specific variant allele, play an essential role. In the case of TP53, this notion has been experimentally addressed by comparing tumor patterns in mice from seven different inbred strains, each having the same R172H *trp53* mutant allele (mouse counterpart of human R175H) in the heterozygous form. This study uncovered significant differences in penetrance, age of onset and tissue type of tumors across different inbred strains, suggesting that the genetic ancestry or background had a clear influence on these phenotypes ^19^. In the case of the Brazilian R337H allele, the genetic founder effect began several hundred years ago and expanded over several centuries of population growth in Southern Brazil, resulting in a population in which about one in three hundred individuals nowadays carry a heterozygous R337H variant on a Lusitanian/Hispanic haplotype of European background ^9,20,21^. There is indeed evidence that the majority of Brazilian R337H carriers are of Lusitanian origin ^20^. Nevertheless, since the population of Southern Brazil is composed of successive waves of migration from Europe, Asia and Africa over a background of native Amerindian populations, the genomes of these carriers are likely to reflect these influences, which is an uncontrolled variable in the population under study. This genetic diversity may represent an important variable to explain why a proportion of R337H carriers may develop cancer only as adults or even remain cancer-free over their entire lifespan.

Sex is part of the genetic makeup of an individual and this study clearly shows that the sex of the patient is in itself a risk factor, with R337H females having a significantly higher risk of developing cancers (65%) than their male counterparts (30%) early in life. Unlike spontaneous cancers where males have a higher incidence of cancers, in LFS Tp53 mutant carriers, females have a higher incidence of cancers at earlier ages than males. This is largely due to the fact that breast tumors are the most common tumor in LFS, independent of the mutant allele. On the other hand males with R337H alleles develop more soft tissue sarcomas ,at all ages, than their females counterparts.

In conjunction to the factors discussed above, multiple other factors may modulate the phenotypes caused by a genetic predisposition. The environment, through its genetic and epigenetic effects, may impact individual risk trajectories in relation with age and cumulative exposure to exogenous risk factors. Immunological response to the tumor and to the variant itself may also be an important factor in determining the lifetime risk ^22^.

Finally there are certainly stochastic events occurring over a lifetime that may contribute to the outcomes of a genetic predisposition to cancer ^19^. Understanding the relative importance of these factors, and their interactions, in determining individual cancer risk, is the focus of molecular epidemiology and is a daunting task in human populations where all these variables can show tremendous variations. A unique genetic predisposition context associated with a founder allele such as R337H may provide an unprecedented setup to further dissect these influences and to better understand the genetic program that leads to the formation of a tumor, its tissue specificity, age of onset, sex, and projected outcomes. Such studies will considerably contribute to a better understanding of the properties that enable stem cells to evolve towards early cancer in a p53-haploid functional or haplo-insufficient environment, and to the mechanisms that underlie the continuous occurrence of such cancers over a lifetime. In the meantime, our analysis of cancer patterns in carriers of the R337H TP53 founder variant will help doctors, genetic counselors, patients and families to elaborate appropriate strategies for increasing awareness, informing and providing adequate early detection measures to the thousands of adult carriers who are at risk in the Brazilian population.

### Research in context Evidence before this study

Since 2001, the TP53 variant R337H (c.1010G>A; p.Arg337His) has frequently been reported in the germline of children with adrenocortical carcinoma and adults in Southern Brazil, with an estimated prevalence of 0.3% in the local population. The magnitude of cancer risk associated with this variant has been a subject of debate: Some carriers exhibit the full spectrum of Li-Fraumeni Syndrome (LFS) traits, while others present with attenuated LFS features or remain cancer-free throughout their lives. However, data on the cancer risk and tumor profiles in adult R337H carriers are scarce, limited to studies with few samples or participants.

### Added value of this study

In this study, we present the Brazilian Li-Fraumeni Syndrome Study (BLiSS), the largest cohort of Brazilian LFS adults to date. It includes 303 Brazilian R337H carriers from 160 families and 42 carriers of non-R337H pathogenic or likely pathogenic (P/LP) TP53 variants from 29 families. This work provides the first systematic, detailed assessment of the tumor spectrum, lifetime cancer risk, temporal trends of tumors, and sex differences associated with R337H. It also compares these aspects with those of carriers of other P/LP TP53 variants. We demonstrate that adult R337H carriers have a significant lifetime risk of multiple cancers that align with the Li-Fraumeni spectrum. However, compared to other P/LP variants, the risk patterns in R337H carriers only partially mirror the LFS spectrum. This study further identifies significant differences in the age of onset and types of tumors between R337H and non-R337H carriers. Notably, we also reveal marked differences between male and female R337H carriers, with females facing a significantly higher risk of developing cancers, predominantly breast cancer, early in life.

### Implications of all the available evidence

Collectively, our findings suggest that the cancer risk associated with the R337H variant in adults is heterogeneous and relies on specific structural, functional, and context-dependent variables that drive its distinct tumor phenotype. Given the high prevalence of R337H in the Brazilian population, our results are poised to aid healthcare professionals, genetic counselors, patients, and families in devising suitable strategies for increasing awareness, providing information, and implementing adequate early detection measures for the numerous adult carriers of the R337H variant at risk in the population.

## Data Availability

All data produced in the present study are available upon reasonable request to the authors

https://docs.google.com/document/d/1LRqXxHgTbkTLN1hvW6iMdxxPyY4YzZlwdxWprWE-TQI/edit?usp=sharing

