## Supplementary material for "Tumor Spectrum and Temporal Cancer Trends in adult carriers of Li-Fraumeni syndrome: Implications for Personalized Screening Strategies in *TP53* R337H carriers"

1 Centro de Oncologia Molecular, Hospital Sirio-Libanes, Sao Paulo, Brazil
2 Department of Oncology, Hospital Sírio-Libanês, São Paulo, Brazil
3 Department of Oncology, Hospital Sírio-Libanês, Brasilia, Brazil
4 Oncologia D'Or, Fortaleza, Brazil
5 Hospital Israelita Albert Einstein, São Paulo, Brazil
6 Serviço de Genética Médica, Hospital de Clínicas de Porto Alegre, Brazil
7 Hospital Universitário Cassiano Antonio de Moraes HUCAM-EBSERH, Espirito Santo, Brazil
8 University Grenoble Alpes, Inserm 1209 CNRS 5309, Institute for Advanced Biosciences, Grenoble, France
9 Institute for Advanced Study, Princeton, New Jersey, USA
*These authors contributed equally to this work
Corresponding authors:

Keywords: Li-Fraumeni Syndrome; R337H variant, *TP53*, germline.

### SUPPLEMENTARY FIGURES

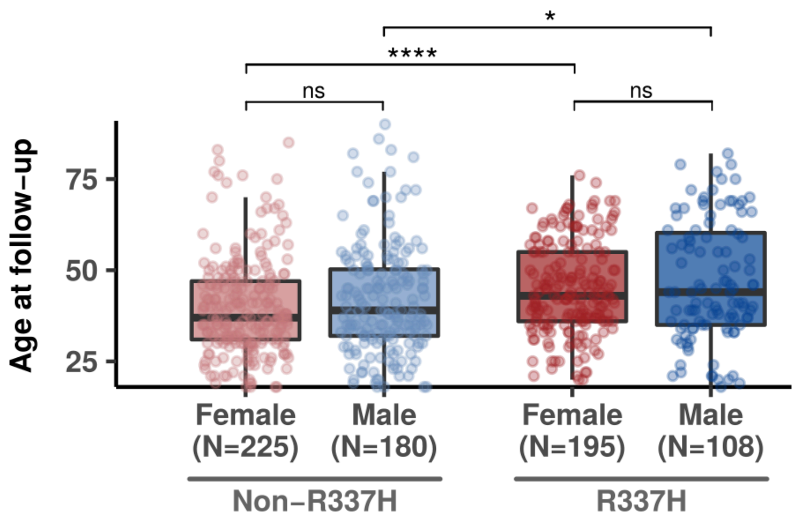

Figure S1. Ages at last follow-up of male and female individuals carrying *TP53* R337H or non-R337H variants (Wilcoxon tests; p-values: **** < 0.0001, * < 0.05, ns “not significant”).

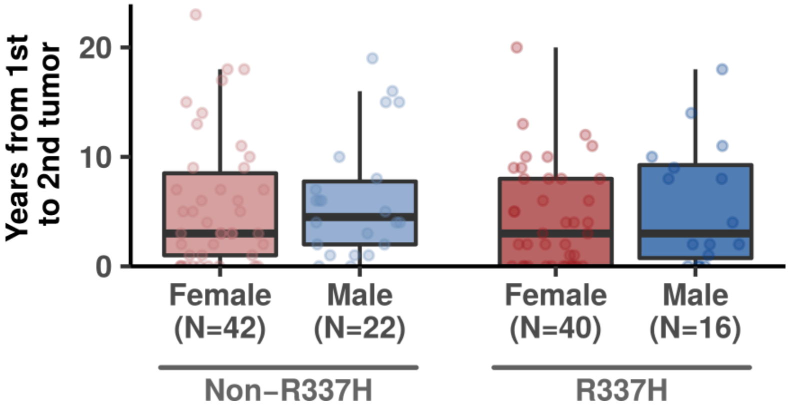


Figure S2. Time elapsed between the first and second diagnosis in male and female individuals carrying *TP53* R337H or non-R337H variants (pairwise Wilcoxon tests: no significant differences among all groups).
